# NEMoE: A nutrition aware regularized mixture of experts model addressing diet-cohort heterogeneity of gut microbiota in Parkinson’s disease

**DOI:** 10.1101/2021.11.10.21266194

**Authors:** Xiangnan Xu, Michal Lubomski, Andrew J. Holmes, Carolyn M. Sue, Ryan L. Davis, Samuel Muller, Jean Y.H. Yang

**Author notes:** Co-last authors (Samuel Muller, Jean Y.H. Yang). Equal contributors (Michal Lubomski, Andrew J. Holmes).

## Abstract

The microbiome plays a fundamental role in human health and diet is one of the strongest modulators of the gut microbiome. However, interactions between microbiota and host health are complex and diverse. Understanding the interplay between diet, the microbiome and health state could enable the design of personalized intervention strategies and improve the health and wellbeing of affected individuals. A common approach to this is to divide the study population into smaller cohorts based on dietary preferences in the hope of identifying specific microbial signatures. However, classification of patients based solely on diet is unlikely to reflect the microbiome-host health relationship or the taxonomic microbiome makeup. To this end, we present a novel approach, the **N**utrition-**E**cotype **M**ixture **o**f **E**xperts (NEMoE) model, for establishing associations between gut microbiota and health state that accounts for diet-specific cohort variability using a regularized mixture of experts model framework with an integrated parameter sharing strategy to ensure data driven diet-cohort identification consistency across taxonomic levels. The success of our approach was demonstrated through a series of simulation studies, in which NEMoE showed robustness with regard to parameter selection and varying degrees of data heterogeneity. Further application to real-world microbiome data from a Parkinson’s disease cohort revealed that NEMoE is capable of not only improving predictive performance for Parkinson’s Disease but also for identifying diet-specific microbiome markers of disease. Our results indicate that NEMoE can be used to uncover diet-specific relationships between nutritional-ecotype and patient health and to contextualize precision nutrition for different diseases.

## Introduction

The human body is home to complex microbial communities, collectively known as the microbiome, which is mostly made up of prokaryotes (bacteria) and archaea ^1^. Considerable evidence has emerged indicating that the microbiome is an important contributor to an individual’s health ^2^. This has been illustrated by links between gut microbiome and numerous diseases, including irritable bowel syndrome ^3^, Crohn’s disease ^4^, type 2 diabetes ^5^, cardiovascular disease ^6^ and Parkinson’s disease (PD) ^7^. The gut microbiome is known to change throughout our lives as a result of various environmental influences. Diet, being one of these factors, has the greatest known long-term interaction with the gut microbiome ^8^. Thus, a deep understanding of the relationship between diet and the gut microbiome and the consequential impact on disease processes, holds promise for developing personalised dietary intervention strategies to modulate and maintain a healthy microbiome population ^9,10^.

Diet has a direct impact on the microbial community in the gut, which governs the activity of the intestinal ecosystem and can have considerable implications for an individual’s health ^11,12^. This is conceptualised in Fig 1 where, for illustration purposes, the macronutrient intake is separated into three perfectly distinct subcohorts with different association between microbiome composition and PD. In practice, several studies have demonstrated that variations in nutrient intake, such as different ratios of protein, carbohydrate ^13^ or dietary fiber ^14^ intake, can influence the host-microbiome association. These discoveries are generally based on elaborate experimental design using model organisms ^13^ or dietary interventions ^15–17^. Recent observational studies suggest that long-term diets could be associated with the microbiome ^18^, and this can further affect overall health. In a similar context, our recent study of the gut microbiome in PD showed that when partitioning individuals based on carbohydrate intake, the predictive performance of the microbiota profile to indicate PD was increased ^19^. Together, these studies suggest that dietary differences can impact relationships between microbiome composition and host health/disease status^27^.

**Fig 1.**
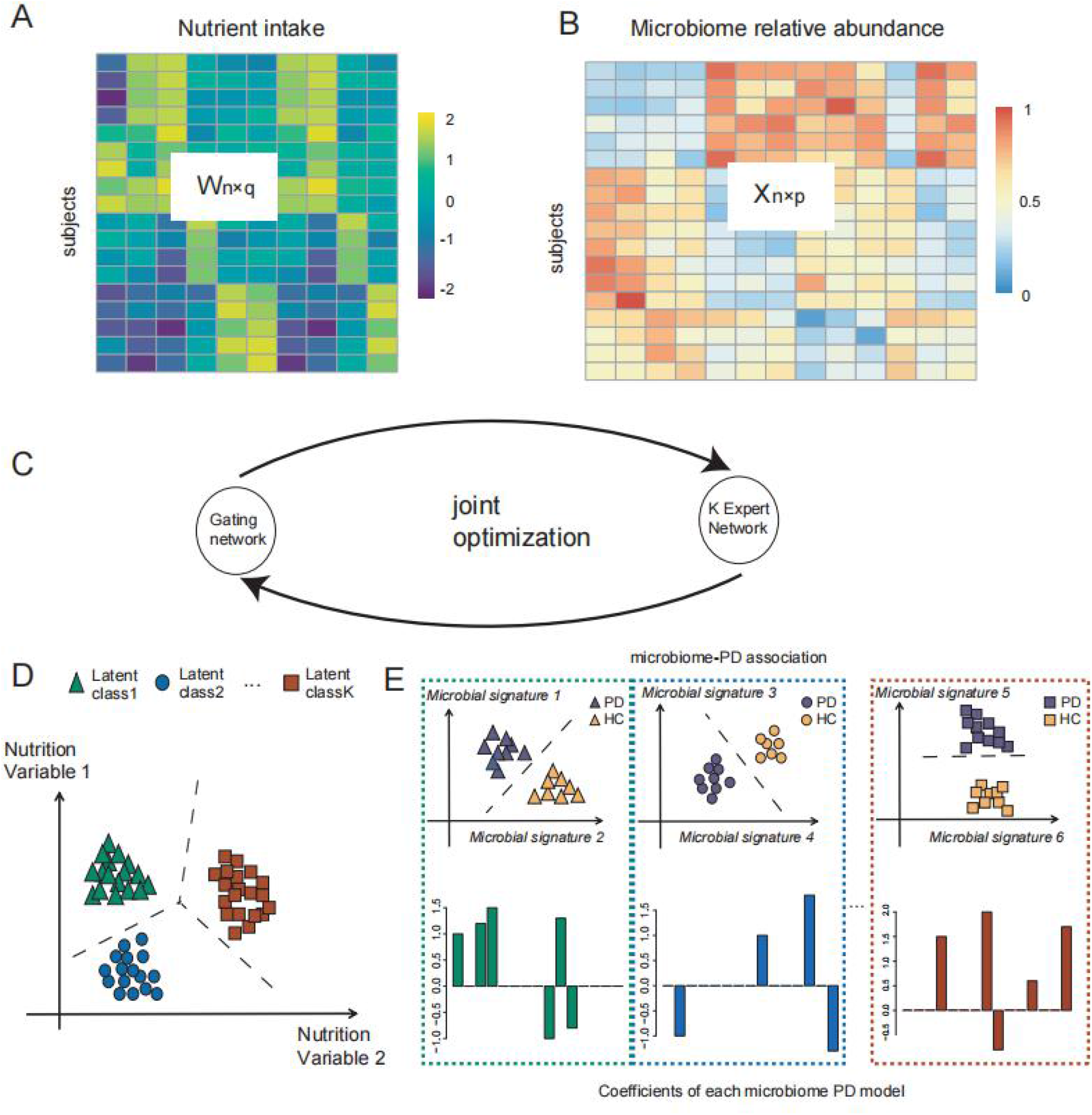
Illustration of NEMoE. **A)-B)** The input matrix of NEMoE: *n* samples with *q* nutrient features and *p* microbiome features **C)** A conceptual workflow of NEMoE, where the joint optimization is achieved by EM algorithm to maximize the regularized likelihood function. **D)** A toy example showing a nutritional-ecotype in the microbiome PD study. The nutrient intake is clustered into *K* latent classes **E)** In each latent class, the microbial signatures of PD are different, which is reflected by the coefficients in the experts network.

To uncover complex heterogeneous relationship structure between diet, microbiome and host health, it is important to identify homogeneous subcohort or latent structure in data that can be explained by a set of features. This is similar to the concept of “ecotype”, which is commonly used to refer to a variant which has observable phenotypically difference in a local environment^20^. Hence, using a data-driven approach, it is able to divide a population into multiple subcohort with distinct microbiological signatures for health that can be best described by nutrient combinations, resulting in what we term “nutritional-ecotypes”. These subcohort can be thought of as diet-based latent classes where they capture interaction between the constraints imposed by nutrient intake of individuals on the community dynamics of their microbiomes ^21,22^.

Methods to discover such diet-based latent classes could be hypothesis-driven based on prior knowledge ^23,24^ or guided by an unsupervised statistical learning method, such as clustering ^25^, followed by latent class analysis ^26^. Although these methods identify nutrient-classes with an altered overall nutritional profile, one limitation is that the defined cohorts may not reflect the heterogeneous microbiome-host health relationship: the drivers of “diet x microbiome” outcomes, “diet x host” outcomes and “host x microbiome outcomes” are overlapping, but not perfectly congruent. Consequently classification models built within a subcohort defined just by diet (or microbiome) will not necessarily improve prediction of the health/disease state ^27^. Similar concepts of identifying cohort heterogeneity to improve prediction performance have been developed in other omics settings and for other diseases ^28,29^. However, simple adaptations of methodologies developed for other omics platforms remain challenging as these do not account for the hierarchical taxonomic structure observed in the study of the diet-microbiome-host interaction. That is, each individual should be in the same diet-specific cohort across all taxonomic levels to keep hierarchical fidelity of the microbial community, i.e. a consistent nutrition class across Phylum, Class, Family Genus *etc*.

To this end, we propose a novel **N**utritional-**E**cotype **M**ixture **o**f **E**xperts (**NEMoE**) approach for uncovering associations between the gut microbiome profile and the health state of an individual that takes into account diet-specific cohort heterogeneity (Fig 1 and Supplementary Fig S1, S2). This is achieved by using a regularized mixture of experts model to simultaneously optimize the separations between nutritional-ecotypes, classification performance of microbiota and the health state. NEMoE also integrates a model parameter sharing strategy to account for the taxonomic information contained in microbiome data, ensuring coherent nutritional classification is maintained across all taxonomic levels. We show through empirical computational simulation research that NEMoE is robust to parameter changes. We also apply NEMoE to real microbiome data from a PD cohort and show that the model outperforms existing approaches of predictive performance and is able to uncover candidate diet-specific microbiome markers of complex disease.

## Results

### NEMoE, a novel method for jointly identifying nutritional-ecotype and for modelling the relationship between microbiota and health state

NEMoE identifies nutritional-ecotypes that represent differential dietary intake as well as the relationship between microbiome structure and host health (Fig 1 and Supplementary Fig S1). This approach has two distinct components: first, a gating network aimed at estimating latent classes shaped by nutritional intake, and second, an experts network aimed at modelling the relationship between the microbiota composition and the health state within each latent class ^30,31^. The input of the gating network is a nutrition matrix, with each variable being the nutrients intake of the individual and the corresponding microbiome measurements are used as input of the experts network. Similar to non-regularized mixture of experts (MoE) models, fitting NEMoE involves estimating the parameters via maximum likelihood estimation to simultaneously optimize the separations amongst nutritional-ecotypes, microbiome classification performance and the health state (Supplementary Fig S2). The optimization procedure is usually achieved by an expectation maximization (EM) algorithm. However, the MoE model does not extend to a large number of feature variables (*p*) and small sample size (*n*) framework, which often occurs in diet and microbiome data where there are many more features than observations. Instead, NEMoE adopts a regularization component to the MoE (RMoE ^32^) by adding elastic net penalties ^33^ on both the gating function and the experts network (details in the Methods section). Next, NEMoE employs a parameter sharing strategy that involves a shared gating network for the microbiome relative abundance matrices across taxonomic levels, to ensure coherent latent classes across all taxonomic levels. Compared with a latent class using purely nutritional intake, our nutritional-ecotype has two advantages: 1) it takes the relationship between microbiome and health outcome into account and is beneficial for identifying diet-specific microbial signatures (Supplementary Fig S1). 2) It incorporates the taxonomic structure in the latent class and keeps hierarchical fidelity of the microbial community.

### NEMoE is able to accurately identify nutritional latent classes shared across different taxonomic levels

We evaluated the efficiency of NEMoE in determining nutritional-ecotypes based on microbiota across different taxonomic levels using both simulated and experimental data. In our simulation study, we created a four-level dataset of 500 samples with shared latent structure, where each individual belonged to a nutritional-ecotype and the relationship between microbiota and health status differed between two simulated nutritional-ecotypes. The adjusted rand index (ARI), a cluster comparison statistic, was used to compare the estimated nutritional-ecotype and the true latent nutritional-ecotype (Fig 2A). We discovered that by incorporating hierarchical taxonomy information in our NEMoE approach, the estimated nutritional class was cohesive and performed better (higher ARI = 0.80) than nutritional-ecotypes estimated from a single taxonomy level (ARI = 0.75). NEMoE achieved this by sharing information across taxonomic levels and the estimated latent class incorporated information from all levels.

**Fig 2.**
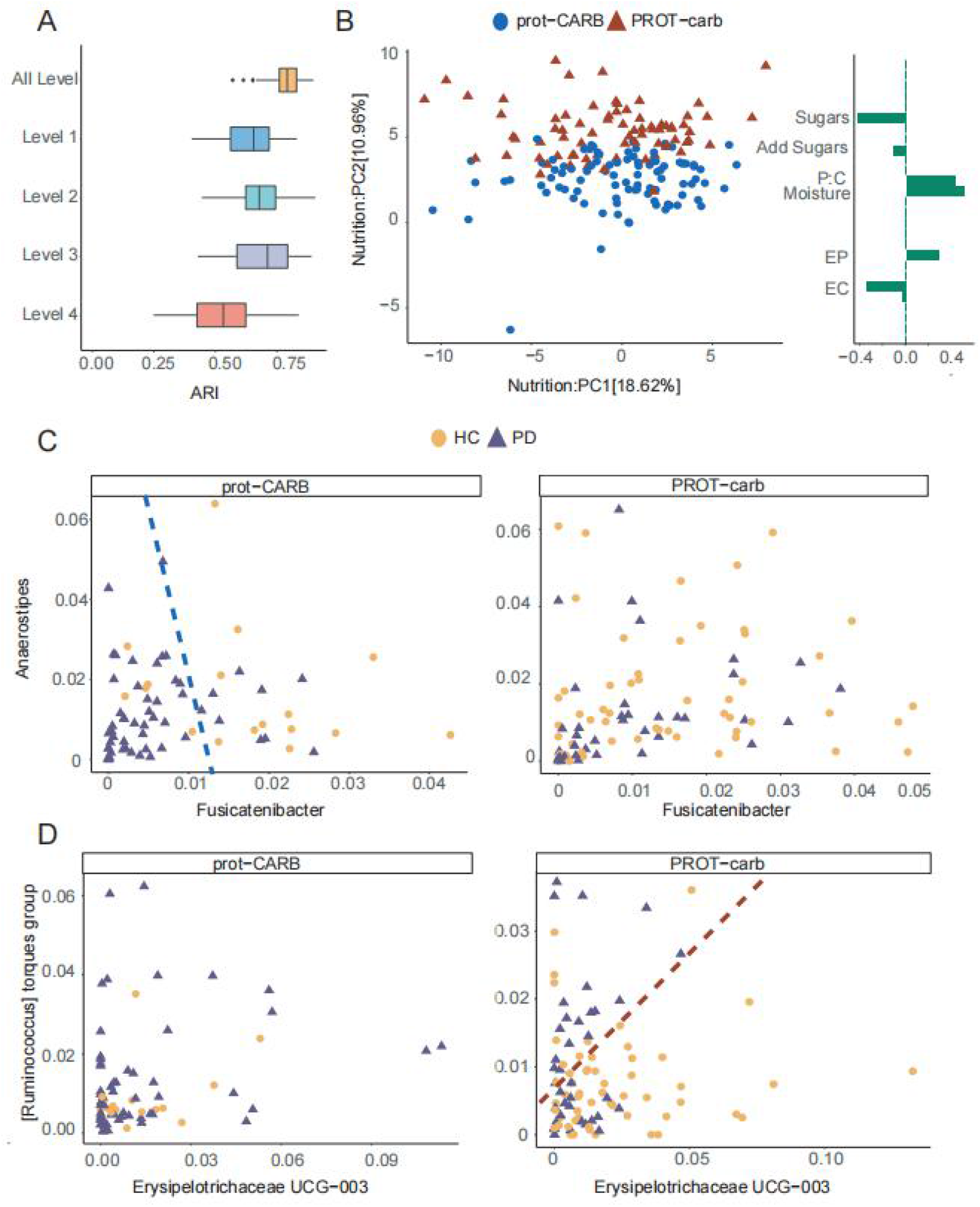
Identification of nutritional-ecotype by NEMoE. **A)** Boxplot comparing NEMoE and single level NEMoE in estimating shared latent classes. The ARI (x-axis) is calculated by comparing the estimated latent class and the true latent class from the data generating model. In all settings, NEMoE using multiple level information performs better. **B)** PCA plot of scaled nutrient intake for subjects colored by the two nutrition classes as estimated by NEMoE. Estimated coefficients of the gating network showed high coefficients for Sugar, Protein:Carbohydrate and Moisture. We denote the two nutrition classes as prot-CARB and PROT- carb with low protein-high carbohydrate intake and vice versa. **C)** Scatter plot of genera *Fusicatenibacter* and *Anaerostipes*. Left panel shows that Parkinson’s Disease and Healthy Controls in the prot-CARB subcohort roughly separate but there is no such separation in the PROT-carb right panel. **D)** Scatter plot of genera *Erysipelotrichaceae UCG-003* and *[Ruminococcus] torques group* showed a different relationship between Parkinson’s Disease and Healthy Controls in two nutritional-ecotypes.

Next, we applied NEMoE to our in-house data from a gut microbiome PD study ^19^. A scatter plot from the first two components of a principal component analysis (PCA) of scaled nutrient intake (details of nutritional features see Methods section) from all individuals is shown in Fig 2B, with the two nutritional-ecotypes best described as “high protein”-”low carbohydrate” (PROT-carb; shown in red) and “low protein”-”high carbohydrate” (prot-CARB; blue). The corresponding loadings show that these two ecotypes have very different ratios of protein to carbohydrate intake. Fig 2C and 2D illustrate that the relationships between gut microbiota and PD status are different between these two nutrition-ecotypes, PROT-carb and prot-CARB. It is important to note that two identified subcohorts are significantly different to clusters identified by unsupervised clustering, such as subcohorts estimated by the kmeans algorithm (ARI ∼ 0, Supplementary Fig S3).

We further established the generalizability of NEMoE by examining its impact when applied to data with different levels of heterogeneity. Here, we created synthetic datasets with four different degrees of separation (Fig 3A, 3B and Methods Simulation studies) and demonstrated that NEMoE performs better than other existing approaches in detecting latent classes and this difference was more evident in challenging situations where the true separation between latent classes was small (Fig S4). This implies that NEMoE has potential to perform well in many observational studies where nutrient intake patterns are mixed or difficult to separate, and hence the NEMoE approach can be applied broadly to human disease datasets with diverse dietary intake.

**Fig 3.**
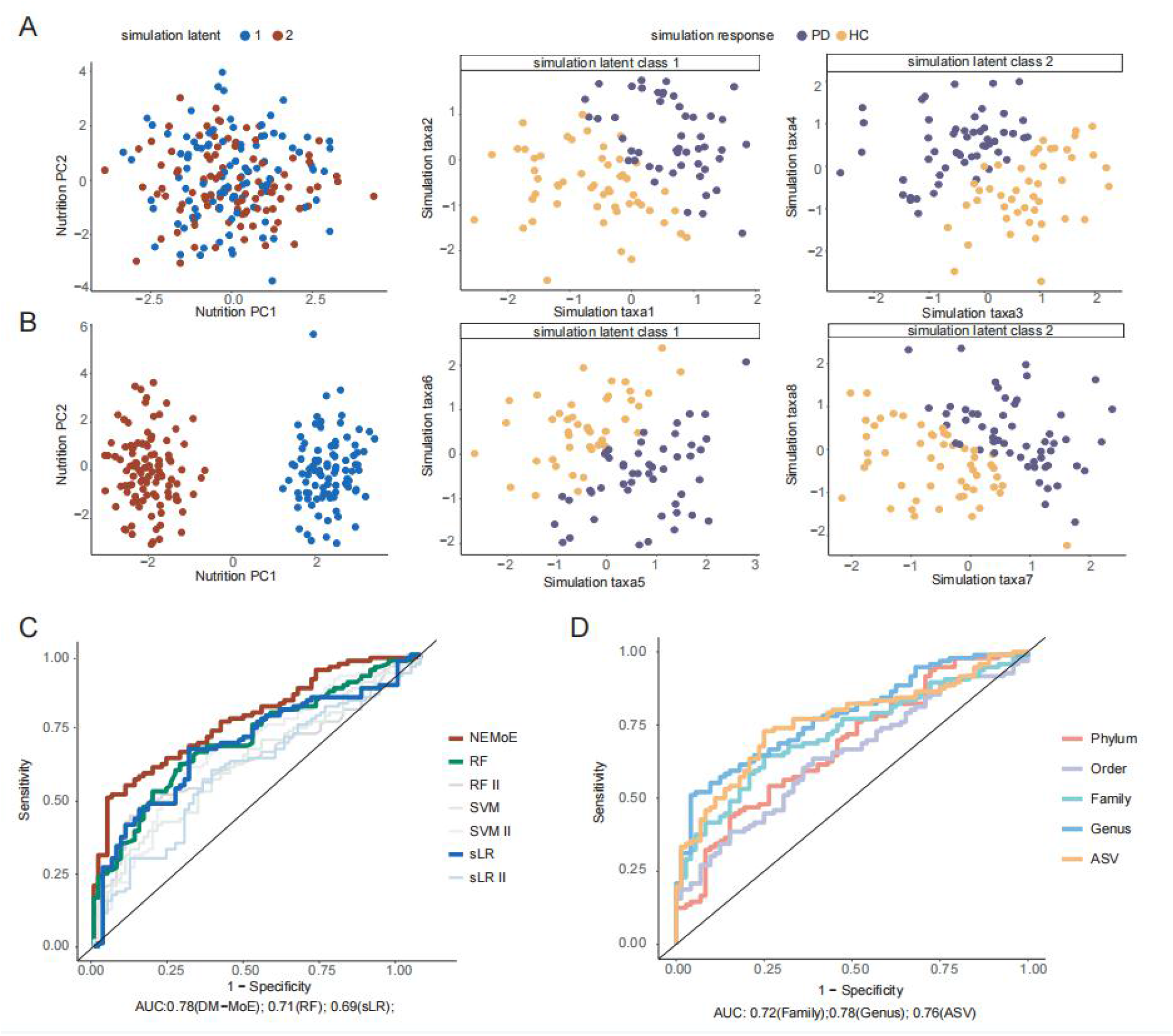
Comparison of NEMoE on simulation dataset and real dataset. **A)** An illustration of a non-separable case where nutrition intake does not show a difference between two nutritional-ecotypes, but each subcohort shows a different relationship between microbiome taxa and health state. **B)** An illustration of a separable case where nutrition intake is significantly different between two nutritional-ecotypes and relationships in each model are similar to the illustration in A). Simulation studies showed that NEMoE can identify both case A) and case B). **C)** Receiver Operating Characteristics curve of different methods (See table S1) in predicting Parkinson’s Disease using LOOCV. NEMoE showed the best LOOCV-AUC (AUC = 0.78). **D)** ROC plot of NEMoE at different taxonomic levels using LOOCV. Genus level showed the best predictive performance(AUC = 0.78).

### NEMoE outperforms existing supervised methods in predicting Parkinson’s disease state

We evaluated the predictive performance of NEMoE using both simulation and real data based on leave-one-out cross validation (LOOCV; see Methods) to the area under the receiver operating characteristics curve (AUC) for the various models described in Supplementary Table S1. In simulation studies we showed that under all comparison settings, NEMoE was able to achieve higher prediction accuracy (Fig S4), which implies NEMoE is robust to different parameter settings, such as *n* and *p*. Fig 3C highlights that when NEMoE was applied to our in-house dataset from a gut microbiome PD study ^19^ with 2 latent classes (AUC = 0.78), it outperformed all other approaches, with the next best being random forest (AUC = 0.71). Fig S6 further highlights that increasing the number of latent classes for this data did not improve the overall AUC.

NEMoE’s ability to detect meaningful subcohorts via its joint optimization approach is a key driver of this increase in accuracy. For example, when comparing to a naive two-stage model that uses unsupervised clustering to identify latent classes before fitting two independent models, the performance of NEMoE is considerably better, as indicated by the large difference in AUC (NEMoE = 0.78, sLR II= 0.6). We further assessed NEMoE’s capabilities on enterotype-separated subcohorts^34^ within our PD dataset. Enterotype, a widely used concept in microbiome research, refers to the categorization of an individual’s microbiomes by the variance in composition ^2,35^. It is widely accepted that enterotype captures stable compositional features of individuals and differences in community-type prevalence across populations with different long-term diets. In this study, we classify 87 samples as Enterotype B, 81 samples as Enterotype F and no samples as Enterotype P. The cluster memberships between the subcohorts determined by NEMoE and by enterotype had no more overlap than pure chance (ARI = 0). Furthermore, building a different classifier for each of the two enterotypes had a much lower (LOOCV-AUC = 0.65) predictive ability than NEMoE (LOOCV-AUC = 0.78). This suggests that NEMoE allows the model to focus more on each latent class and increases prediction performance by more precisely identifying subcohorts with differential microbiome-PD relationships.

### NEMoE is able to identify informative taxonomic levels and consensus candidate microbial PD signatures in multiple independent cohorts

In our in-house gut microbiome PD investigation, NEMoE provided a natural criterion to examine which of the five taxonomic levels (Phylum, Order, Family, Genus, and ASV) was most informative with respect to different nutrient intake. We achieved this by evaluating predictive performance for PD at each taxonomic level to determine the most informative. Fig 3C shows that genus was most predictive compared to the other taxonomic levels, with an LOOCV-AUC of 0.78.

Next, our NEMoE model determined a separate set of PD microbial signatures for each nutritional-ecotype. The derived coefficients represent the level of association between microbiota and health/disease state (e.g. PD) in each nutritional-ecotype (Fig 4A and 4B) and results for all taxa are given in Additional Data 1. We can broadly group the microbiota taxa into five categories based on their coefficient estimates: (i) significant in both classes with different directions; (ii) significant in both classes with the same direction; (iii) significant in prot-CARB only, (iv) significant in PROT-carb only and (v) not-significant in both classes. The first category “significant in both classes with different directions” represents consistent abundance changes in both nutritional-ecotypes (Fig 4B). It was noted that the genera *Fusicatenibacter* and *Blautia* showed consistent negative coefficients in both PROT-carb and CARB-prot nutritional-ecotypes. Such genera may be considered stable PD microbial signatures, with several studies showing their underrepresentation in PD^36–40^.

**Fig 4.**
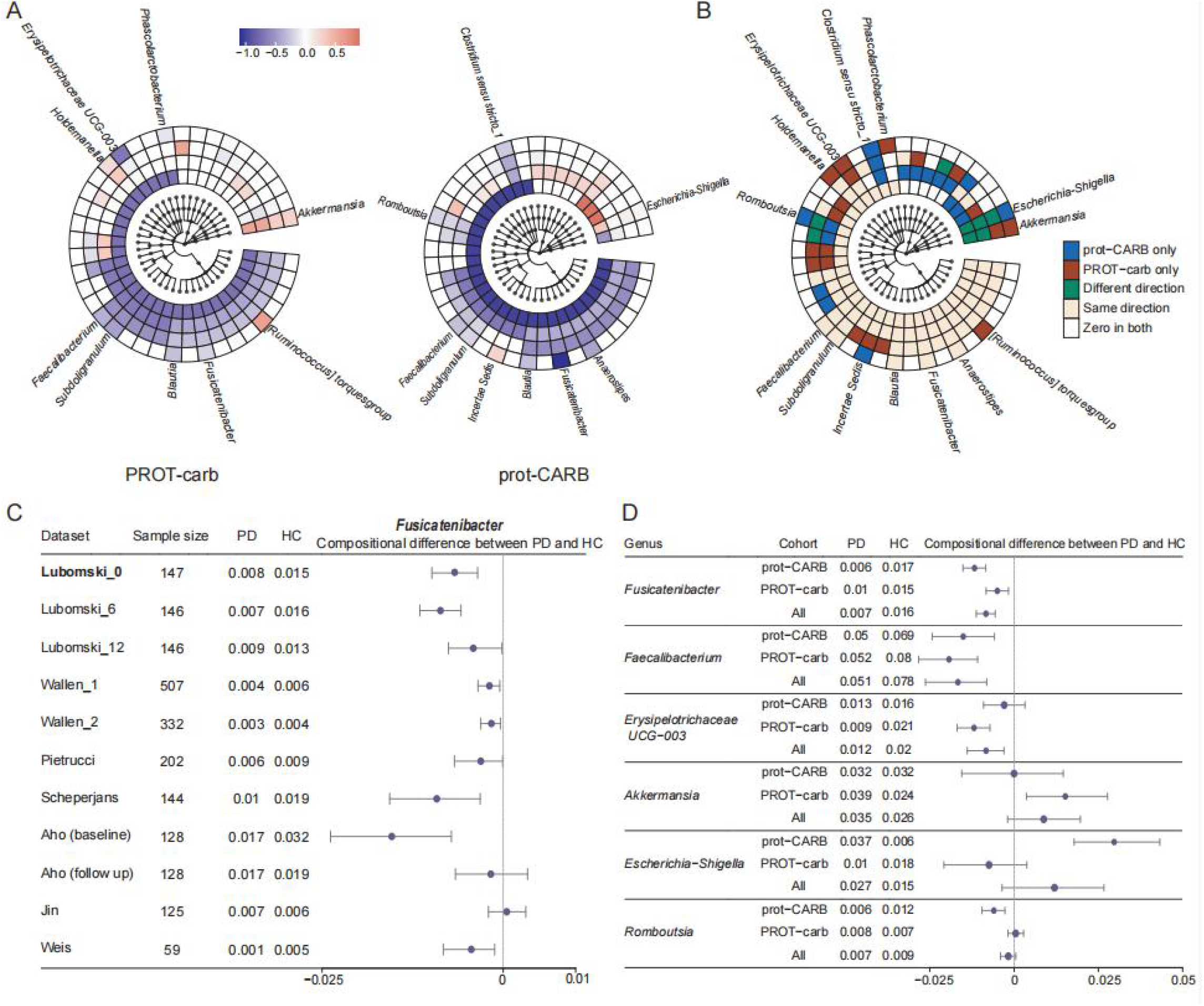
Results of NEMoE on gut microbiome-PD study. **A)** Coefficients of experts network in NEMoE at different taxonomic levels. The two latent classes showed distinctly different microbiome patterns. **B)** Identification of diet specific microbial signatures of PD. The “Same direction*”* class showed consistent function in different dietary patterns. The “PROT-carb only” and “CARB-prot only” classes tended to be important only with specific dietary intake. The “Different direction” class changed their coefficients in different dietary patterns. **C)** Validation of differential relative abundance of genus *Fusicatenibacter* in 11 different datasets. With the exception of one dataset (Jin *et al* 44) all other datasets showed decreasing *Fusicatenibacter* in PD. **D)** Forest plot of 95% confidence interval of selected taxa showed NEMoE is able to identify the species that are differentially represented in specific nutritional-ecotypes.

The underrepresentation of *Fusicatenibacter* and *Blautia* were further validated using data from eight independent PD microbiome studies (Table S2). We processed the publicly available datasets using the dada2 pipeline ^41^(v1.16) and taxonomy reference “*silva* 138” ^42,43^. The relative abundance changes of the genus *Fusicatenibacter* were examined across all datasets, as shown in Fig 4C. In all but one dataset ^44^, *Fusicatenibacter* had significantly lower relative abundance among PD individuals. Similar results were observed for *Blautia* (Fig S5), verifying NEMoE’s ability to identify consensus microbial signatures of PD in multiple independent cohorts.

### NEMoE is able to identify the species that are differentially represented in specific nutritional classes

We note that taxa categories (i)-(iii) represent differential abundance changes that are unique in the two nutritional-ecotypes prot-CARB and PROT-carb, which indicate some microbial signatures of PD are diet-specific (Fig 4C). We discovered that the genus *Escherichia-Shigella* was significantly underrepresented in the prot-CARB nutritional-ecotype but not in the PROT-carb ecotype. This genus belongs to the family *Enterobacteriaceae* (including *E. coli, Shigella, Salmonella, Klebsiella*, etc.), which are facultative anaerobes and known for utilizing soluble sugars as a carbon source. When an individual’s diet has a higher intake of sugars (or simple starch) it can be expected that the relative abundance of these microbiota will likely increase. Recent studies found that *Escherichia-Shigella* is a pathogenic bacteria that potentially reduces short-chain fatty acid production and produces endotoxins and neurotoxins ^45,46^.

We also found a significant increase in the relative abundance of the genus *Akkermansia*, but only in the PROT-carb class (Fig4D). These bacteria are known to impact immune response and constipation, with many studies reporting an overrepresentation in PD ^37,38,40,47^. *Akkermansia* breaks down mucins and turns them into short-chain fatty acids; further, their relative abundance is thought to increase when “diet-specialize bacteria” decline as a direct impact of changes in microbially accessible carbohydrates (MAC). Generally, a low carbohydrate diet will lower MAC, thus lowering the number of diet-specialist microbes and allowing *Akkermansia* to become overrepresented, consistent with our discovery.

Most importantly, neither of these two genera (*Escherichia-Shigella, Akkermansia)* were discovered in our previous analysis using the ALDE model ^48^, where both classes were combined for microbiome biomarker identification (*Escherichia-Shigella:* p-value 0.14, *Akkermansia: p*-value 0.55) ^19^. This highlights the relevance and importance of nutritional-ecotypes identification in microbiome marker discovery.

## Discussion

The aim of this study is to investigate and unravel the complex interaction between diet, the microbiome and an individual’s health. We achieve this by exploring the effects of dietary pattern (or composition) on the relationship between the microbiome and host health and by developing a method called NEMoE that detects such heterogeneity. Through a series of simulation studies, NEMoE shows strong prediction performance when the underlying data show heterogeneity explained by different nutrient intake. Furthermore, we illustrate the practical performance of NEMoE on a gut microbiome PD study in which nutritional-ecotypes and microbial signatures of disease are found. We show that NEMoE outperforms the predictive accuracy of previous models (higher AUC) and identifies multiple known PD microbiome markers. Two different nutritional-ecotypes are also identified within our data with distinct protein-to-carbohydrate intake ratios and novel candidate signatures that were indicative of a diet-specific cohort.

While we focus on discovery of microbial signatures of PD by splitting the population based on dietary profile, the architecture of NEMoE means its flexible algorithm can take different types of data for subcohort detection (data used for gating networks) or biomarker identification (data used for expert networks). Therefore, an alternate research question could be to identify nutrients as disease markers for diverse microbiome profiles, and the NEMoE system can readily adapt to this new problem by changing the input of the gating network and experts network. Often, clinical knowledge or interest guides the decision on question formulation. However, if we consider both the dietary and microbiome profiles to be equivalent proxies for one’s nutrition system, then performing NEMoE in two different ways allows us to empirically compare the effectiveness of nutritional signatures versus microbial signatures and provides us with insight into the natural heterogeneity in the microbiome and in nutritional intake.

NEMoE is designed to partition samples based on their associated nutrient intake and can be viewed as a data-driven strategy for subcohort or latent class identification. An alternative option is to investigate a knowledge-driven strategy to achieve the same goal and one example is the use of “enterotype”. Similar to unsupervised learning, stratifying samples based on “enterotype” whilst providing an alternative way to stratify samples, does not explicitly take disease prediction performance into account. As a result, the aggregate predictive ability of the three separate enterotypes is lower than the nutritional-ecotypes division discovered by the NEMoE approach.

The proposed NEMoE method is based on diet-microbiome-host health interaction. However, it is not restricted to diet and microbiome data. Our method can be expanded to other multi-omics studies to identify subcohorts determined by the heterogeneity in relationships between covariates and response. One potential application is in the clinical heterogeneity of the relationship between multi-omics and host health. In such scenarios, the subcohorts are determined by their clinical index while the omics data are used to model the relationship between host health and information from a specific molecular platform.

In summary, we present NEMoE, a novel statistical method to model heterogeneity of diet and the gut microbiome in disease. NEMoE identifies nutritional-ecotypes based on a maximum likelihood framework and using an Expectation-Maximization step to estimate the model parameters. Our proposed framework also enables identification and then accounts for multiple levels of structure in the feature set, a unique characteristic in microbiome data, where we are able to estimate a shared latent class for each individual at different taxonomic levels. Effectiveness of NEMoE is validated at three levels. First, we demonstrate through a series of extensive simulation studies the model’s ability to accurately identify latent classes and to increase microbiome predictability. Second, we validate the performance of NEMoE on a real disease dataset and show that this method outperforms existing two-stage methods. Finally, the downstream impact and practical importance of NEMoE is further demonstrated by the discovery of diet-specific PD microbiome markers, such as *Escherichia-Shigella* and *Akkermansia*, which are not identified by the ALDE model ^48^.

## Methods

Methods, including statements of data availability and any associated accession codes and references, are available in the online version of the paper.

## Supporting information

External Data

## Data Availability

All data produced are available online at https://sydneybiox.github.io/PD16SData

## Author contributions

JYHY, SM and XX conceived the study. XX led the method development and data analysis with input from SM and JYHY. XX led the evaluation of the method with input from all authors. ML, CMS and RLD provided the case study data and guided evaluation of the method. AH analyzed and interpreted the microbiome results. JYHY, XX and SM wrote the manuscript with input from all co-authors. All authors read and approved the final version of the manuscript.

### Acknowledgements

The authors thank all their colleagues, particularly at The University of Sydney, Sydney Precision Bioinformatics and Charles Perkins Centre for their support and intellectual engagement. Professor Vicki Flood and Jon Flood for assistance analysing and interpreting the Food Frequency Questionnaire data. The authors also sincerely thank all participants for their patience and willingness to contribute to this research.

## Online Methods

### Datasets

#### PD-microbiome

A gut microbiome dataset containing 101 PD patients and 83 healthy controls. The stool samples were collected and 16S rRNA V3-V4 amplicon sequencing was performed on an Illumina MiSeq platform. Ethical approval was granted by the Northern Sydney Local Health District Human Research Ethics Committee and the North Shore Private Hospital ethics committee, HREC/18/HAWKE/109, SPHEC 2018-LNR-009 respectively.

#### PD-diet

Dietary information was collected by a comprehensive Food Frequency Questionnaire and resulted in a table of nutrient intake with 23 macronutrients, presented earlier ^49^. Details of the sample information and sequence processing can be found in Lubomski *et al* ^19^.

#### Public validation (PV) studies

We curated a series of datasets from eight different publicly-available microbiome studies ^7,36,44,50-54^ to further validate results from NEMoE. All the datasets were processed using the dada2 pipeline ^41^ (v1.16) and microbiome taxa were annotated using taxonomy reference “silva 138” ^42,43^. Samples with low sequence reads (<1000) were excluded from the analysis. More information on these datasets can be found in Table S2.

### Data processing

#### PD-microbiome data processing

We excluded 7 samples with extremely large energy intake (>20,000 kJ per day), one subject with low microbial read counts (Total counts < 10000) and two samples with missing nutrition measurements, resulting in 175 samples (75 HC individuals and 100 PD individuals). Raw counts from microbiome data were first normalized by total sum scaling, *i*.*e*. the counts (totals) were normalized into a composition proportion. Then core microbial features were kept and further transformed: Features that had more than 30% zeros in the *n* samples and features which had sample variance smaller than 10^−5^ were filtered out at each taxonomic rank resulting in the core microbial features of 7 Phylum, 19 Order, 27 Family, 41 Genus and 101 ASVs and 3,152,746 total reads were kept from 6,024,011 reads ; variance stability transformation, *i*.*e*. an arcsin square root transformation, was performed on taxa proportion ^55,56^; the arcsin transformed data were further standardized to have mean zero and unit variance (z-score).

#### PD-diet features construction

In addition to the nutrients intake values, we calculated the percentage of energy intake as protein (EP), percentage of energy intake as fat (EF), percentage of energy intake as carbohydrate (EC) and protein intake and carbohydrate intake ratio (P:C) as additional variables. These transformations of nutritional features are widely used in nutri-omics studies ^57,58^. All of the 27 nutritional features were z-scored.

### Nutrition-Ecotype Mixture of Expert (NEMoE) model

The development of NEMoE was inspired by a mixture of experts approach to model heterogeneous data as shown in Fig S2A. In machine learning, the concept of “*gate*” ^59^ can be thought of as a decision-making component given some input. Our approach consists of two key components, a “*gating network*” that is set up to determine which nutritional-class the sample belongs to and a “*k-experts network*” of size *k* to build classifiers for each nutritional-class. NEMoE uses a regularized MoE (RMoE) model, which adds elastic-net penalties to both the gating network and the experts network. Regularization is needed here because a non-regularized MoE does not extend to a large *p* small *n* framework ^60^ where the number of features (*p*) is much larger than the number of samples (*n*). This data characteristic often occurs in diet and microbiome data where there are many more microbial features (*p*) than individual samples (*n*). NEMoE further incorporates the taxonomic information into RMoE by jointly optimizing RMoE models from all taxonomic levels with the added constraint that all RMoE share the same gating network (Fig S2B).

#### Mathematical formulation of NEMoE

For a transformed microbiome data at taxonomic level *l*, we use the matrix 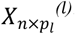 to denote the relative abundance in *n* samples of *p*_*l*_ taxa. The corresponding diet information, measured as a nutrients intake matrix, is denoted as *W*_*n*×*q*_, where the *q* columns are the nutrient metrics for the same n samples Let *Y*_*n*_ denote the binary response of the health outcome, with *Y* = 1 and *Y* = 0 representing individuals with and without disease, respectively. NEMoE models the heterogeneous relationship between the microbiome and the health outcome by a mixture distribution, i.e.

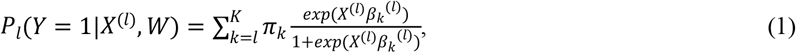

where 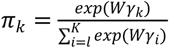 is the nutrition class mixing weight of shared components determined by nutrients intake, and where *γ*_*k*_ and *β*_*k*_ are the corresponding effect size for the gating network and the experts network, respectively, and *K* denotes the predetermined number of nutrition classes.

NEMoE estimates the regularized sum of all levels of the log-likelihood function in Equation (1), where the regularization term consists of elastic net penalties for both the gating network and the experts network:

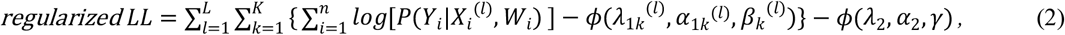

where 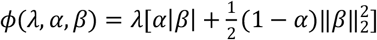 is the elastic net penalty function and *λ*_1*k*_^(*l*)^ *α*_1*k*_^(*l*)^ *λ*_2_ *α*_2_ are the corresponding parameters for penalties in the experts network and in the gating function.

### Model Fitting

NEMoE is fitted based on an Expectation Maximization (EM) algorithm. We explored four different types of initialization methods to alleviate the effect of starting points of the non-convex optimization. Different variants of EM algorithms including ordinary EM, classification EM (CEM), stochastic EM (SEM) and simulated annealing EM (SAEM) ^60,61^ were also implemented to suit different scales of the problem.

The tuning parameters in NEMoE can be selected based on different criteria. Our NEMoE package has implemented selection criteria based on the Akaike information criterion (AIC), Bayesian information criterion (BIC), integrated classification criterion (ICL) and cross validation (CV) ^32,62,63^. A warm-start procedure is used similar to the warm-start in the glmnet package ^64^ to accelerate the evaluation of NEMoE under different parameters. Details of these methods can be found in the reference manual of the NEMoE package https://sydneybiox.github.io/NEMoE.

### Simulation studies

Our simulation is inspired and integrated from multiple simulation studies. These include the generation of the nutrition data based on a multivariate Gaussian distribution ^65^ and a sparse multinomial regression model; the generation of the microbiome data using a zero-inflated latent Dirichlet allocation model (zinLDA) ^66^ and constructing the health outcome using sparse logistic regression ^55^. The details of the simulation are described as follows.

The simulation of the nutritional data consists of two main components, the values of the nutritional measurements (*W*) and the underlying latent class which we refer to as nutritional-ecotype (*Z*).

#### Constructing the effect size between *W* and *Z*

For a given number of nutritional features *q* and given the total number of latent classes *K*, we first simulate the effect size between *W* and *Z*, denoted by the matrix *γ*_*q*×*k*_ where each element *γ*_*jk*_, *j =* 1, …, *q* and *k=* 1, …, *K*, represents the effect size of the *j*^th^ nutrition feature on the *K*^th^ latent class. We randomly select five nutrition features to have non-zero effect size and its value is either *c*_*g*_ or − *c*_*g*_ with equal probability, *i*.*e*.

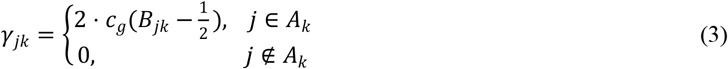

where 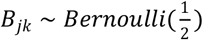 and *A*_*k*_ denotes a set of nutritional variables with non-zero effect size randomly drawn from the *q* nutritional variables, *c*_*g*_ *> 0* is a constant that controls the overall strength of the effect size and *B*_*jk*_ is a Bernoulli random variable with probability parameter 1/2. This is designed such that the effect of a nutritional feature can randomly have a positive or negative value with equal probability. Selection of five nutrition features allows only a fraction of nutrition features to contribute to the ecotype.

#### Simulating nutritional values *W* and nutritional-ecotype *Z*

The nutritional values are simulated with a *q*-variate Gaussian mixture distribution with *K* components. Given the *i*^th^ sample, we first generate its nutritional-ecotype *Z*_*i*_ by randomly drawing an index from the set {1, …, *K*} indicating which latent class it belongs to. For *Z*_*i*_ *= k*, its corresponding nutrition data *W*_*i*_ is generated from a normal distribution

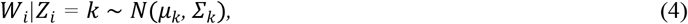

where *μ*_*k*_ and Σ_*k*_ are the mean and covariance matrix of the *k* mixture component, respectively. In our simulation, we set Σ _*k*_ to be a matrix with diagonal entries equal to 1 and off-diagonal entries equal to *ρ*, we further set *μ*_*k*_ to be proportional to the effect size of the *k*^th^ latent class *γ*_*k*_ *i*.*e*.

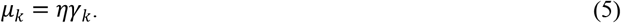

The use of Gaussian mixture distributions for nutrition data is an extension of the simulation method by Chen and colleagues^65^. The levels of heterogeneity between dietary patterns were controlled by different levels of η. A larger η represents a greater gap between the means of the components and thus, the larger that gap the easier it is to distinguish the nutritional ecotype *k* (Fig 3A). In our simulation, we define “none”, “weak”, “mild” and “strong” separation by setting the maximum absolute value η to be 0, 0.1, 0.3, and 0.5, respectively.

#### Simulating microbiome data *X*

The generation of the microbiome data follows a zero-inflated latent Dirichlet allocation model (zinLDA)^66^. This model simulates several typical characteristics of the microbiome data including over-dispersion, zero-inflation and high dimensionality. The distribution of the microbiome data *X* is given by,

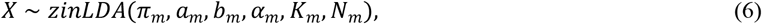

where *N*_*m*_ is the total number of reads, *K*_*m*_ is the number of microbiome subcommunities, α_*m*_ is the parameter in the Dirichlet distribution, *π*_*m*_ relates to the generation of the subcommunity, *a*_*m*_ and *b*_*m*_ are the parameters in the zero-inflated generalized Dirichlet distribution related to the generation of counts within each subcommunity. Our simulation uses the default parameters as in zinLDA^66^: *N*_*m*_ was drawn from a discrete uniform distribution with a lower bound of 5,000 and an upper bound of 25,000, *π*_*m*_ *=* 10, *K*_*m*_ = 5, *π*_*m*_ *=* 0.4, *a*_*m*_ *=* 0.05 and *b*_*m*_ *=* 10, respectively. The microbial counts at ASV level were generated using the corresponding distribution of *p* variables.

For each simulation, we generate *n* samples of microbiome data at 5 taxonomic levels, where level 1 to level 5 correspond to Phylum, Order, Family, Genus and ASV, respectively and (*p*_1_, …, *p*_5_) = (10, 20, 50, 80, *p*) corresponds to the number of microbiome variables for each level respectively. The hierarchical structure of the taxonomic table is generated by randomly grouping microbial features from the higher neighbouring level *l*+1 into the *p*_1_ group when going from the (*l*+1) level to the l level. We denote *ASV*^(*l*)^(*j*)as the set of taxa at ASV level that is mapped to taxa *j* at level *l*. The simulated microbiome data of the *l* taxonomic table was thus hierarchically aggregated from the taxa from the (*l+*1) taxonomic table and the corresponding counts matrix is denoted as 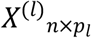.

#### Simulating health outcome *Y*

The relationship between microbiome and health outcome was simulated based on a mixture sparse logistic regression which extends the model by Dong and colleagues ^55^. We simulate the binary health outcome *Y* in three steps.

Step 1: Simulate the microbial signatures of each latent class at ASV level. The microbial signatures were selected from candidates’ taxa that satisfy two conditions: prevalence is larger than 50% (counts are non-zero in at least 50% of samples) and the variance of its relative abundance is larger than 10^−6^. Then we randomly select 5 taxa from the candidates taxa for the *k*^th^ latent class and denote the corresponding sets of taxa as *Ak*^(*ASV*)^, *k = 1*, …, *K*.

Step 2: Simulate the effect size between microbiome *X*^*(l)*^ and health outcome *Y* at the *l*^th^ taxonomic level and latent class *k*. We first generate the effect size at ASV level, which we denote as *β*_*k*_^(*ASV*)^. The *i*^th^ element of *β*_*k*_ ^(*ASV*)^ was generated as follows:

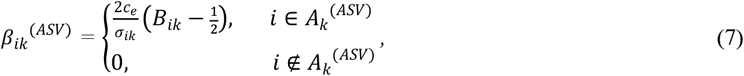

where 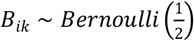. Then the effect size of the *j*^th^ taxa at level *l* (*β*_*jk*_^(*l*)^) is generated by aggregating its corresponding effect size at ASV level,

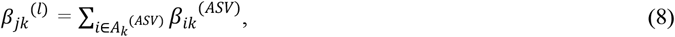

where 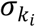 is the standard deviation of taxa *i* in nutrition class *k* and *c*_*e*_ *> 0* is a constant that controls the strength of the effective size, and *B*_*jk*_ is a Bernoulli random variable with probability parameter 1/2.

Note that for taxonomic level *l*, the corresponding effect sizes were generated as the sum of the level *l+1* coefficients similar to as in Wang and colleagues ^67^. Here, the strength of effect size is inversely proportional to 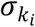 and ensures that the contribution of the microbiome signature is not mainly affected by its relative abundance.

Step 3: Simulate the health outcome *Y*. We simulate the probability that “the health outcome *Y* equals 1” as an average of all 5 levels mixture sparse logistic regression with the mixing weight 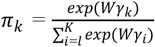,

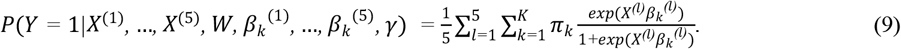

For given microbiome data 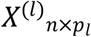, nutrition data *W*_*n*×*q*_ and the corresponding effect size *γ* and *β*_*k*_^(*l*)^, (*l =* 1, …, 5). we draw *n* Bernoulli pseudo-random samples with probability parameter given by equation (9) to obtain the binary health outcome vector *Y*_*n*×1_.

### Performance evaluation

#### Comparison methods

Table S1 contains a summary of all methods used in the comparison study. We included the most commonly used methods in microbiome analysis as well as a naive two-stage approach. All of the comparisons were performed on simulation datasets and on in-house data on the Genus level.

#### Naive two-stage approach

The approach first clustered the nutrition data using unsupervised learning methods such as kmeans. Then, based on the clustering result, samples in each cluster were used to build a classification model of microbiome and health state. The choice of classification models we used in our simulation includes sparse logistic regression, support vector machine and random forest.

#### Differential abundance

We compared differential relative abundance between PD and HC in all datasets. The comparison was based on a non-parametric bootstrapping procedure. We resampled the data with replacement, then calculated the difference of the average relative abundance between PD and HC. This procedure was repeated 10,000 times for each taxa and the 95% confidence interval of the differential relative abundance was calculated.

#### Simulation framework

Our simulation first generated independent data of 2*n* samples from the procedure described above, then the first *n* samples were used for training and another *n* samples were used to calculate the predicted accuracy. The details of parameter settings in each simulation is described in Table S3.

## Data availability

All the data used were published previously and the corresponding information is shown in Table S2. All processed datasets are incorporated in a R data package that is freely available from our GitHub repository at https://sydneybiox.github.io/PD16SData.

## Code availability

NEMoE is implemented using Rcpp and available at https://github.com/SydneyBioX/NEMoE and in the process of submission to the BioConductor repository. All code used in this paper is freely available from our GitHub repository https://github.com/SydneyBioX/NEMoE_MS.

## Supplementary Figures

**Supplementary Fig S1.**
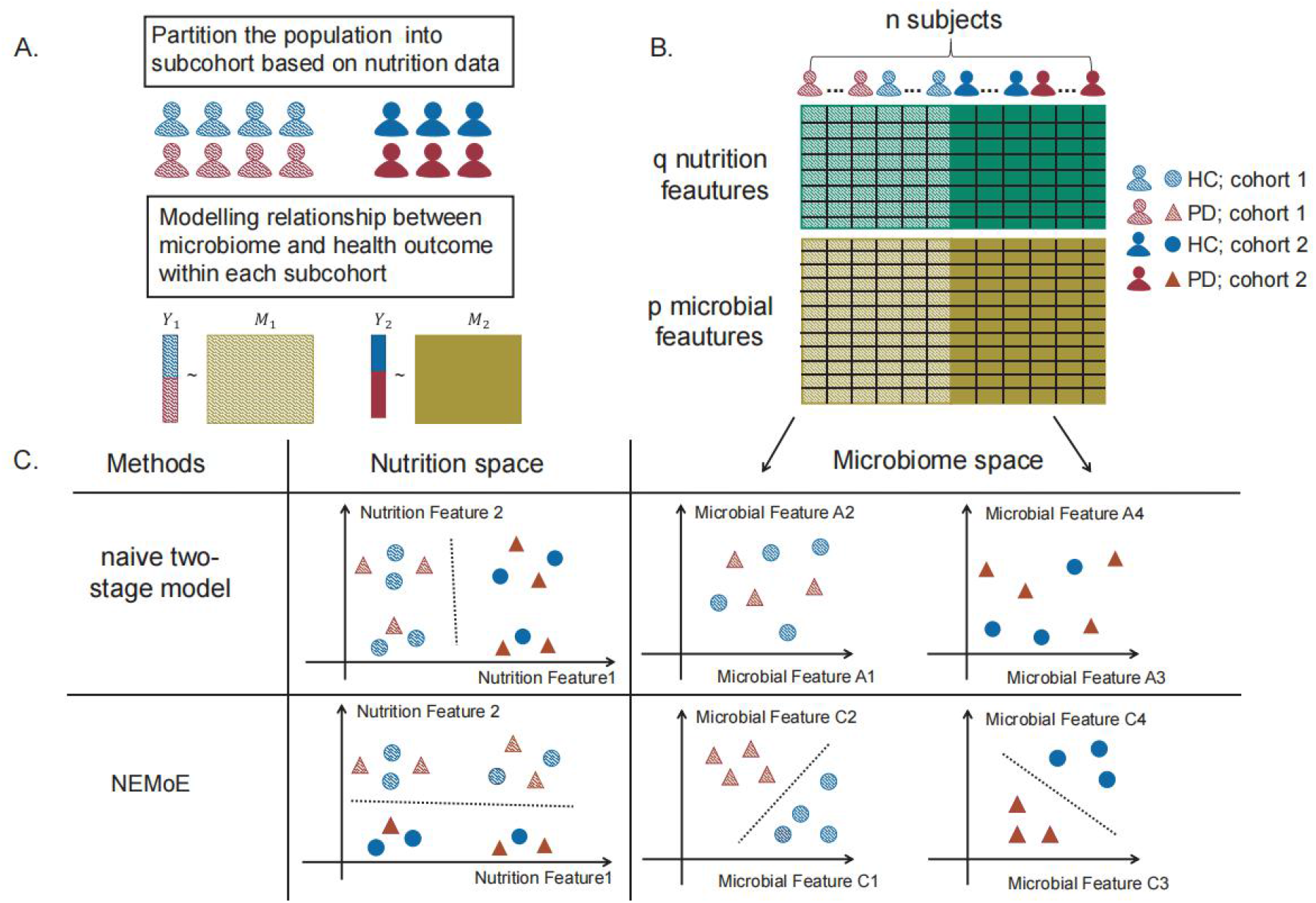
Illustration of NEMoE and two-stage model. **A)** A common workflow of a two-stage model: first clusters the cohort based on the nutrition intake, then builds a model between microbiome data and health outcome within each cohort. **B)** Illustration of a two-stage model with two latent classes. **C)** Illustration of two methods NEMoE and naive two-stage model in both nutrition space and microbiome space. Naive two-stage model identified two latent classes showed best separation in the nutrition space but do not count for the relationship between microbiome and health outcome; Two latent classes identified by NEMoE showed differential relationship between microbial features and health outcome.

**Supplementary Fig S2.**
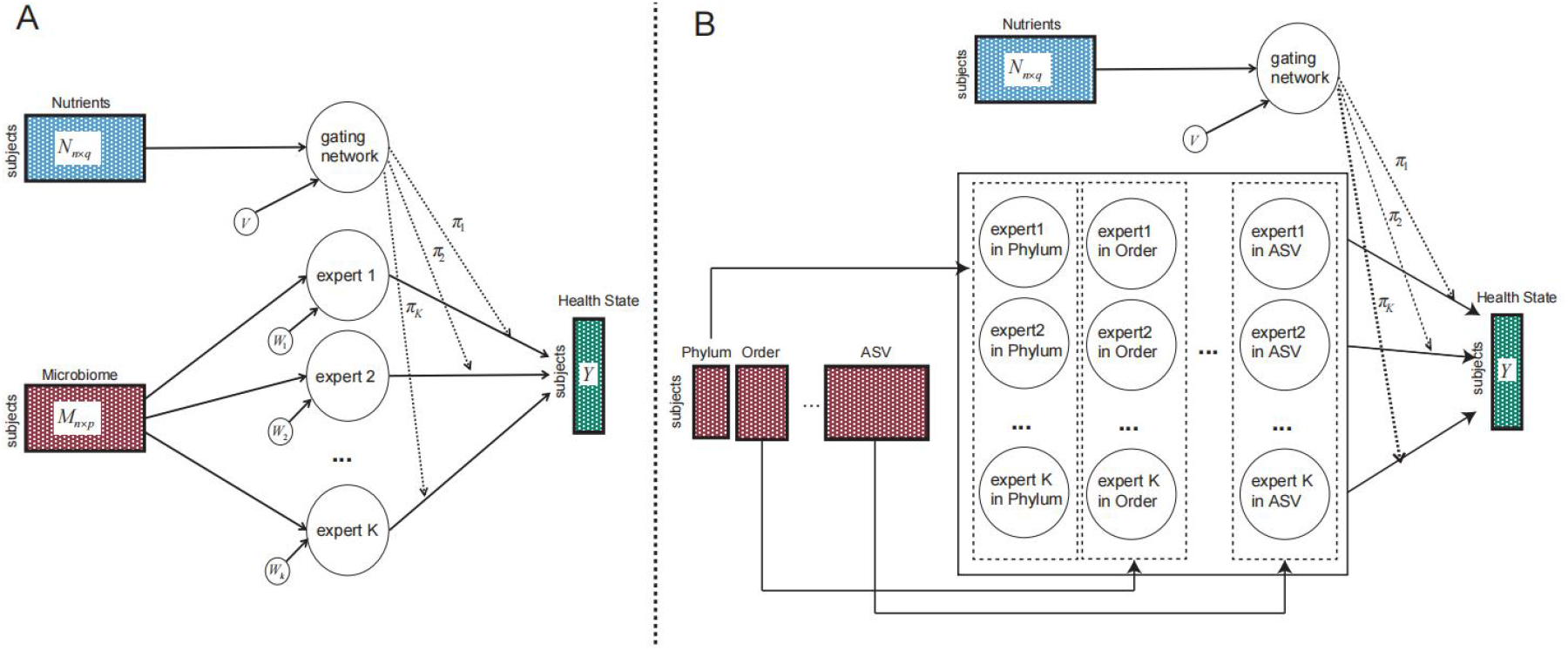
Graphical model representation of NEMoE. **A)** A graphical model representation of the mixture of experts model. **B)** A graphical model representation of the NEMoE extends the MoE to multi-level with a shared gating network.

**Supplementary Fig S3.**
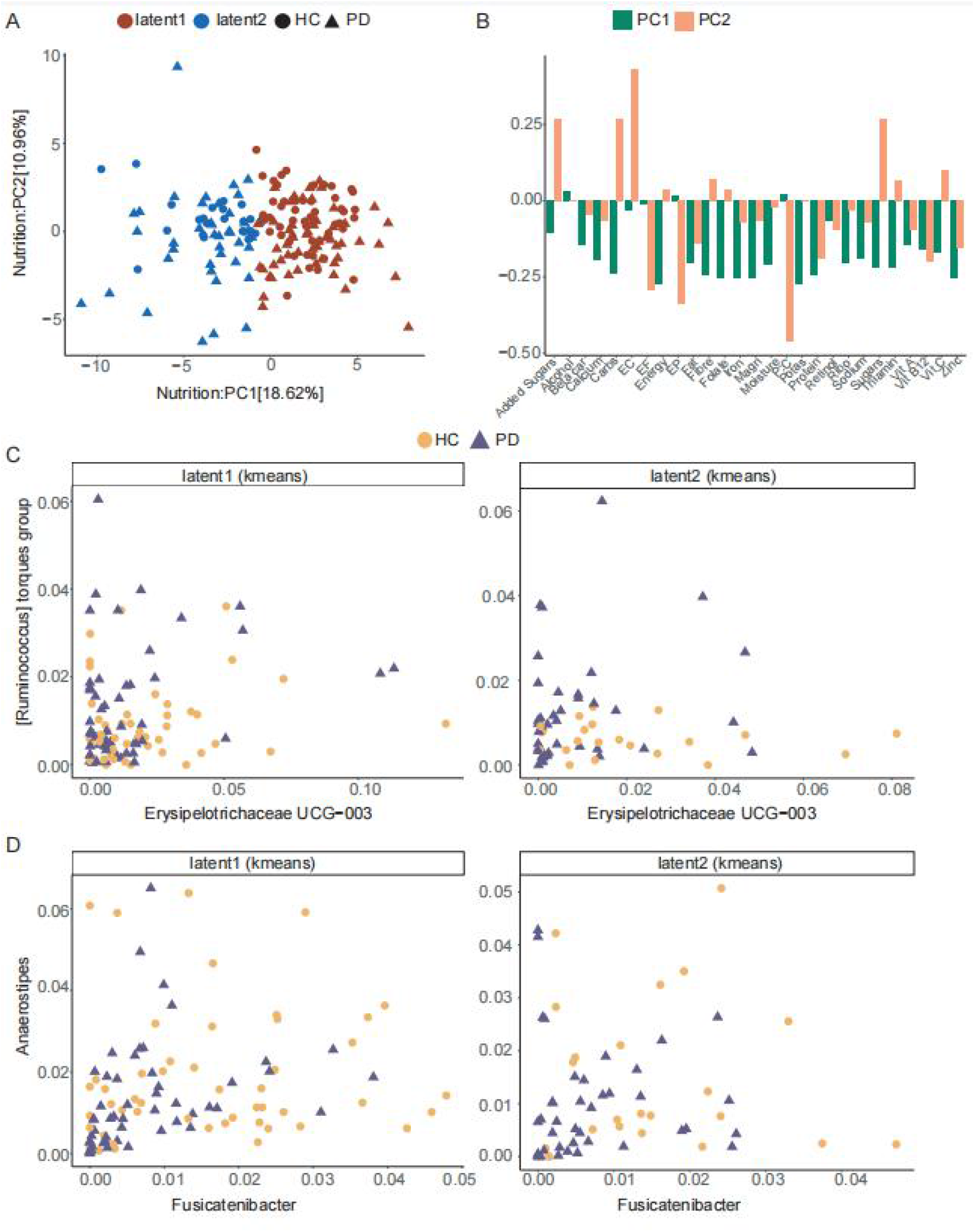
Nutrition classes determined by kmeans do not show an informative relationship between microbiome and PD. **A)** PCA plot of scaled nutrient intake for subjects colored by two latent classes estimated by kmeans. **B)** Loadings of the first two PCs. Loadings of the second PC shared some important variables with the coefficients of the gating network in NEMoE. **C)** and **D)** Variables selected by NEMoE do not show a clear difference between PD and HC.

**Supplementary Fig S4.**
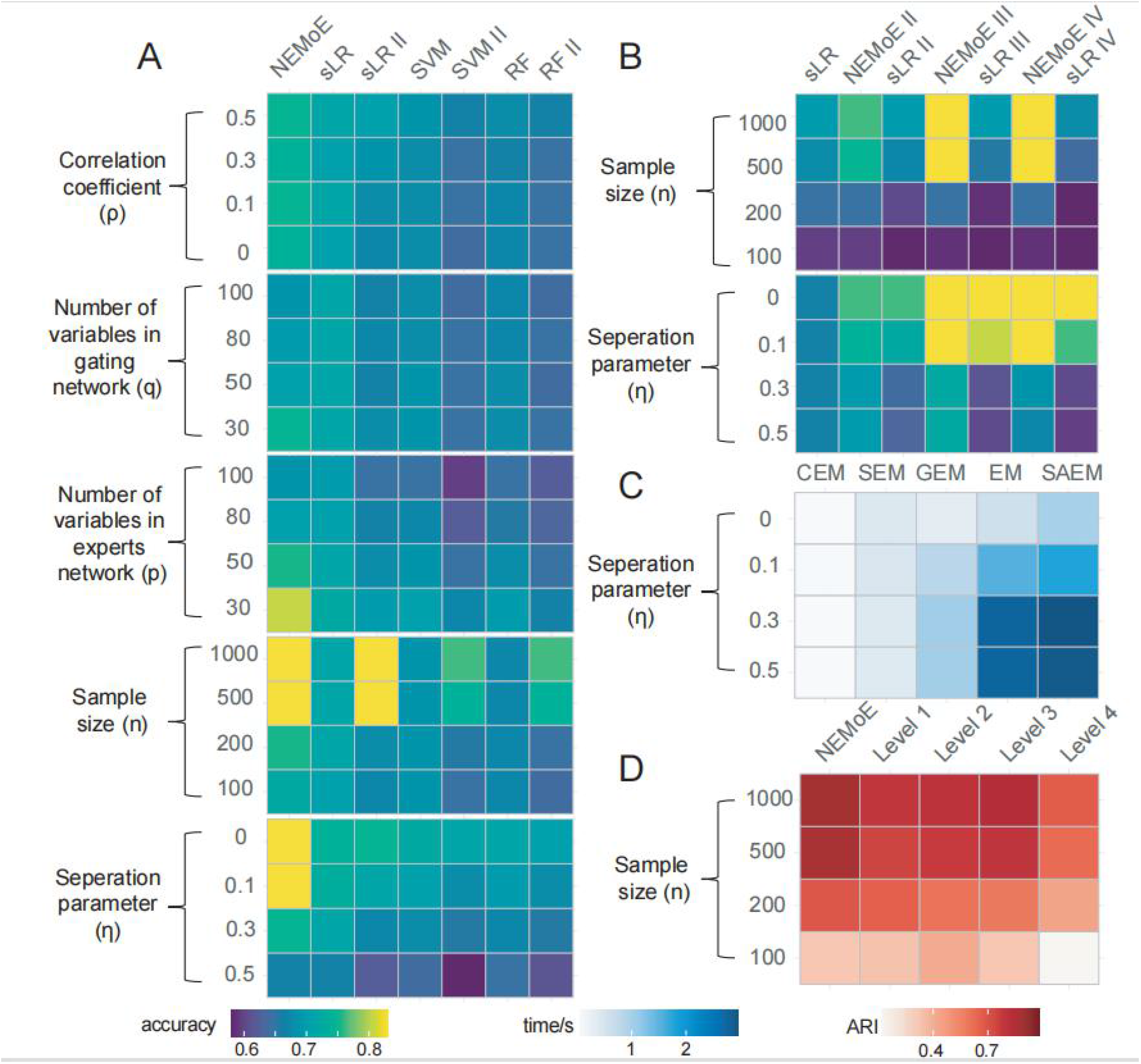
Simulation results of NEMoE and other methods under different settings. **A)** simulation results under different simulation parameters settings including the separation parameter *η*, sample size *n*, number of variables in the gating network *q* and in the experts network *p* and correlation *ρ* between variables. The NEMoE achieves best predictive performance compared with others in all settings with latent class structure. **B)** Simulation results of three latent classes with different *n* and *η*. NEMoE III and NEMoE IV perform well under most parameter settings. **C)** Time consumption of different EM-algorithms implemented in NEMoE. CEM was the fastest while achieving lower regularized LL, while EM and SAEM achieved higher regularized LL but required more time. **D)** Comparison between different taxa levels with NEMoE in estimation of shared latent classes. Level *K* represents fitting RMoE with data in level *K* (*K*=1,2,3,4). The ARI is calculated by comparing the estimated latent class and the generated true latent class.

**Supplementary Fig S5.**
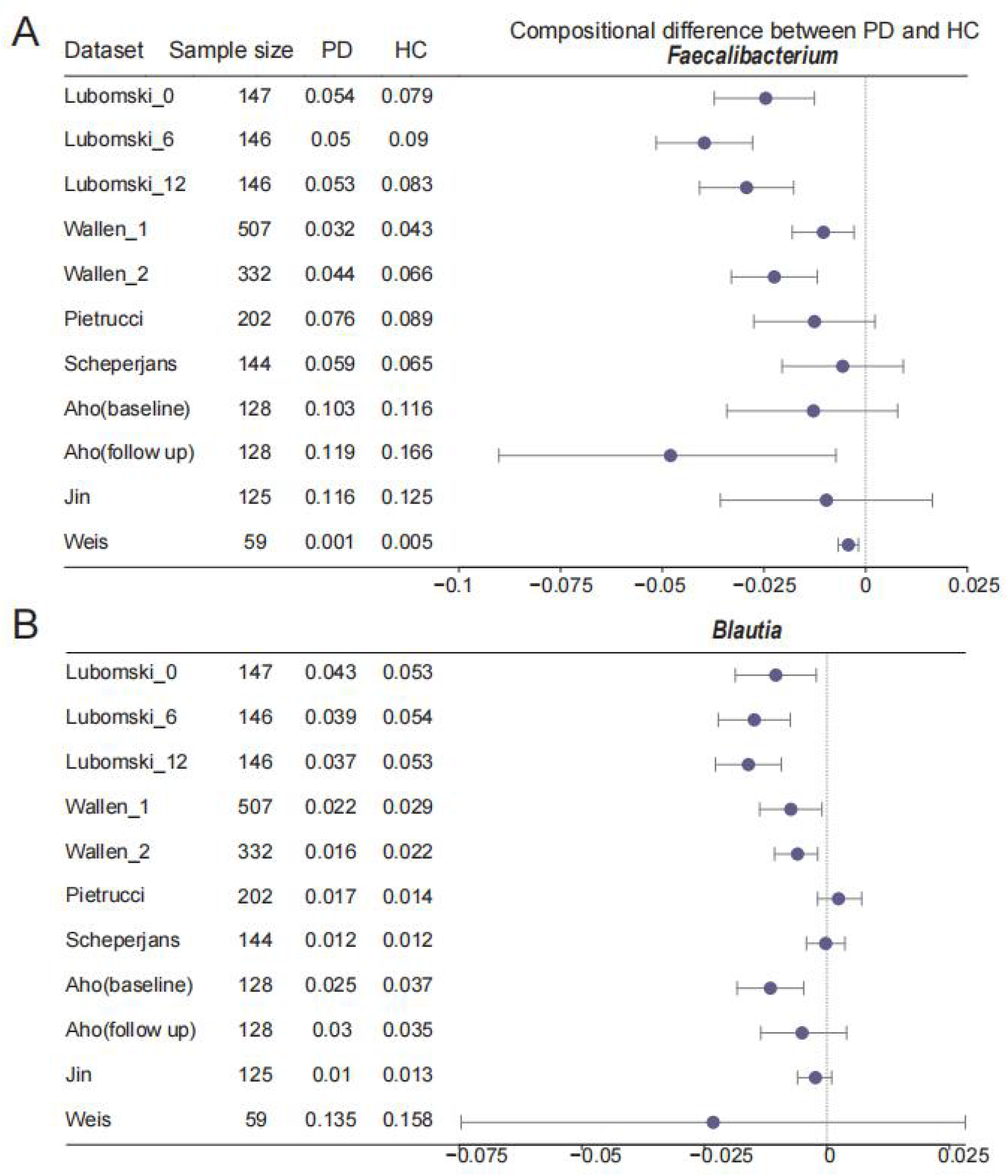
External validation of consensus taxa *Faecalibacterium* and *Blautia*. **A)** Validation of differential relative abundance of genus *Faecalibacterium* in eight different datasets. **B)** Forest plot showing the validation of differential relative abundance of genus *Blautia* in eight different datasets.

**Supplementary Fig S6.**
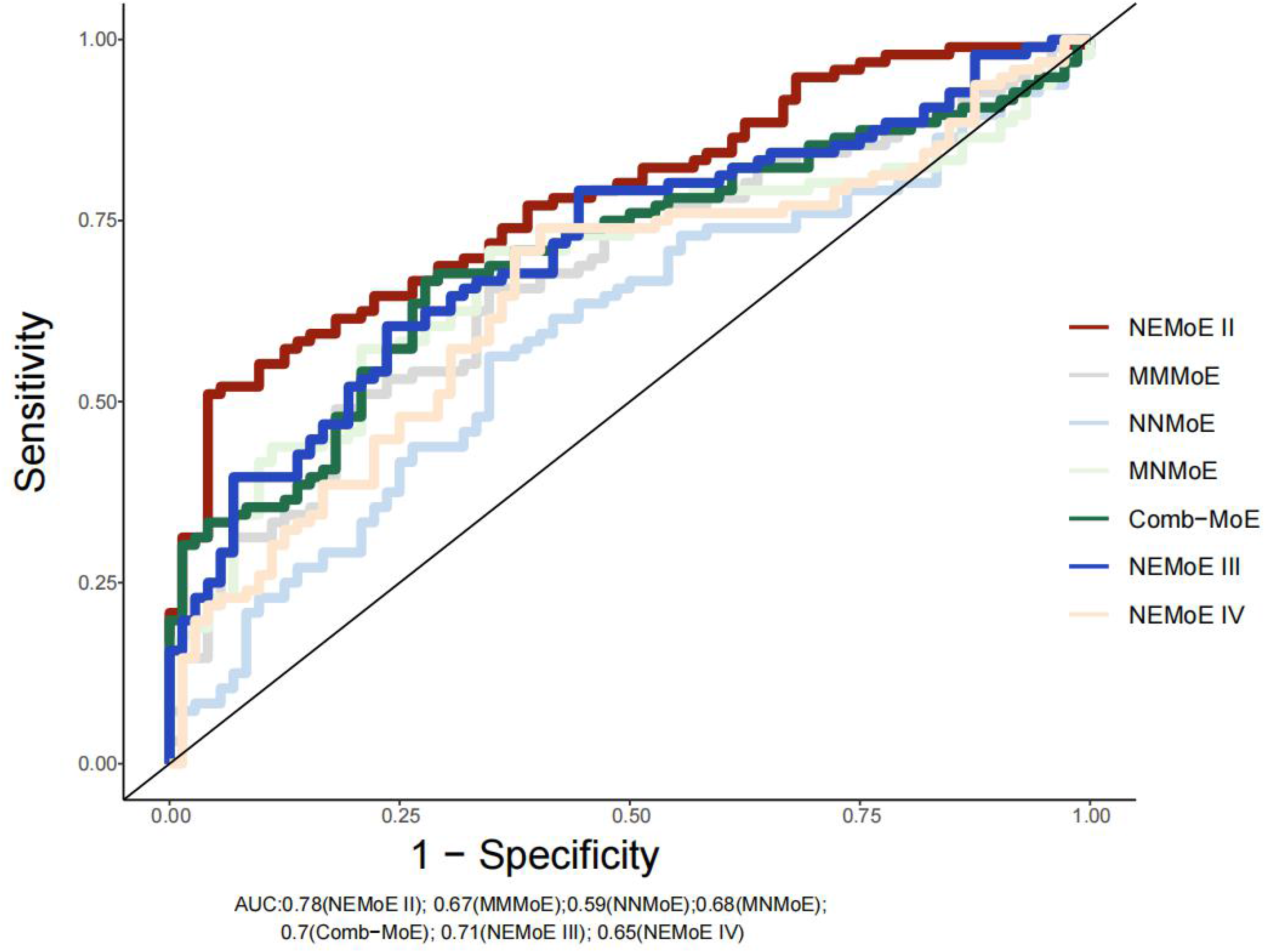
Prediction performance of different types of input for NEMoE. Using nutrients intake as the input of the gating network and microbiome as the input of the experts network showed better prediction performance than for the other cases. NEMoE with 2 nutrition classes showed best prediction performance in our dataset.

## Supplementary Table

**Supplementary Table S1.**
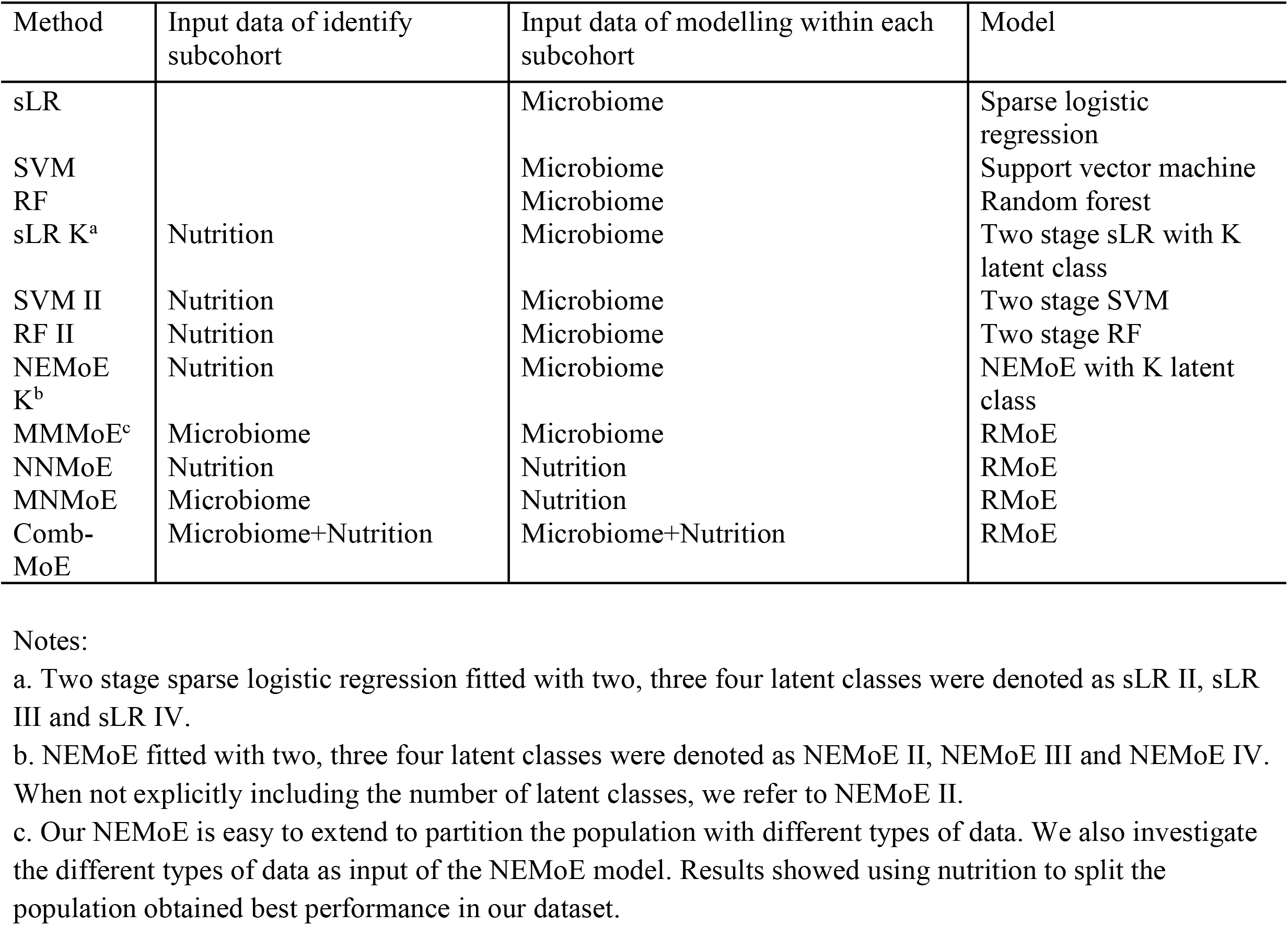
Summary of methods for comparison.

**Supplementary Table S2.**
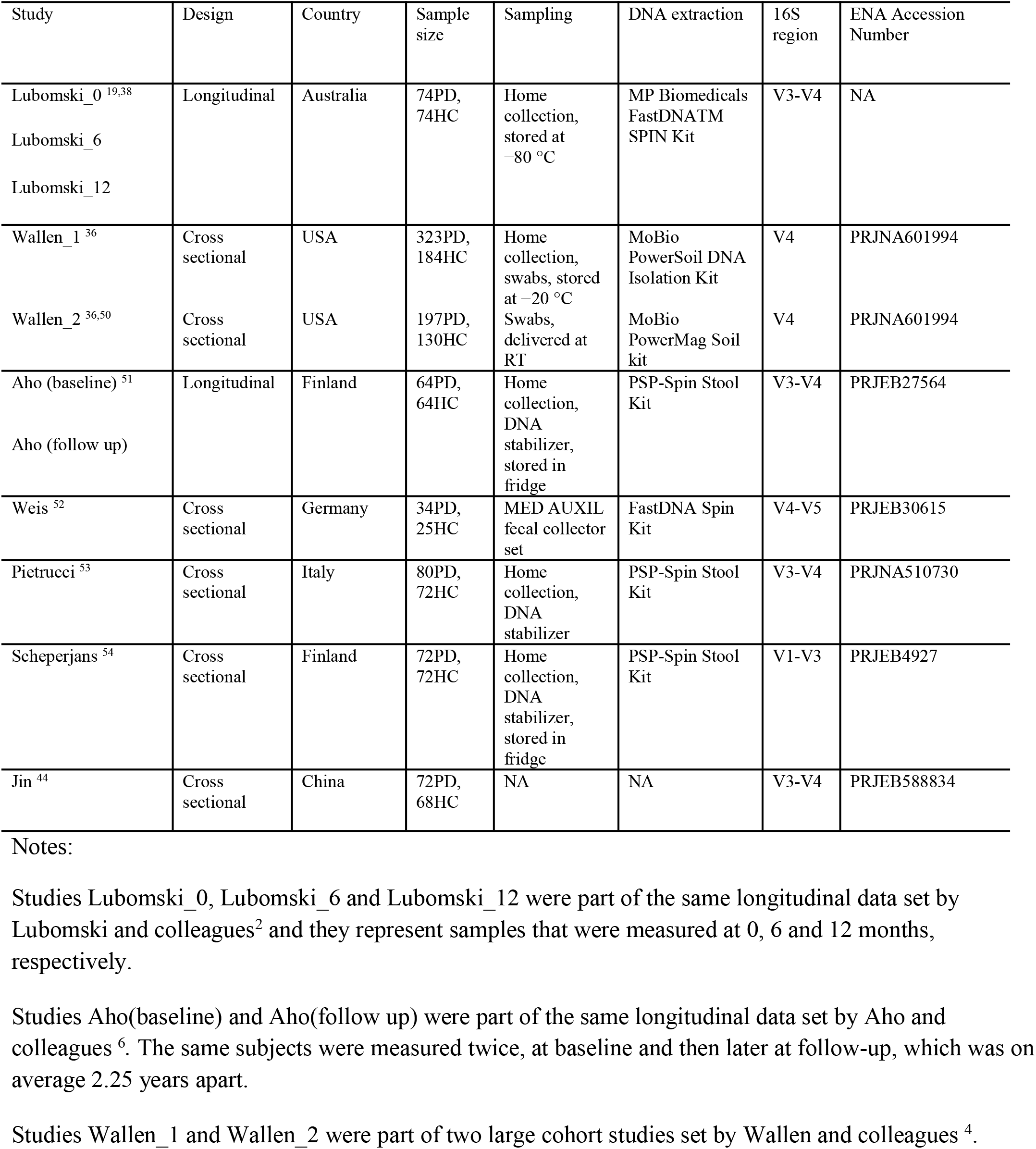
Summary of eight publicly available Parkinson’s Disease microbiome studies used for validation of the NEMoE model.

**Supplementary Table S3.**
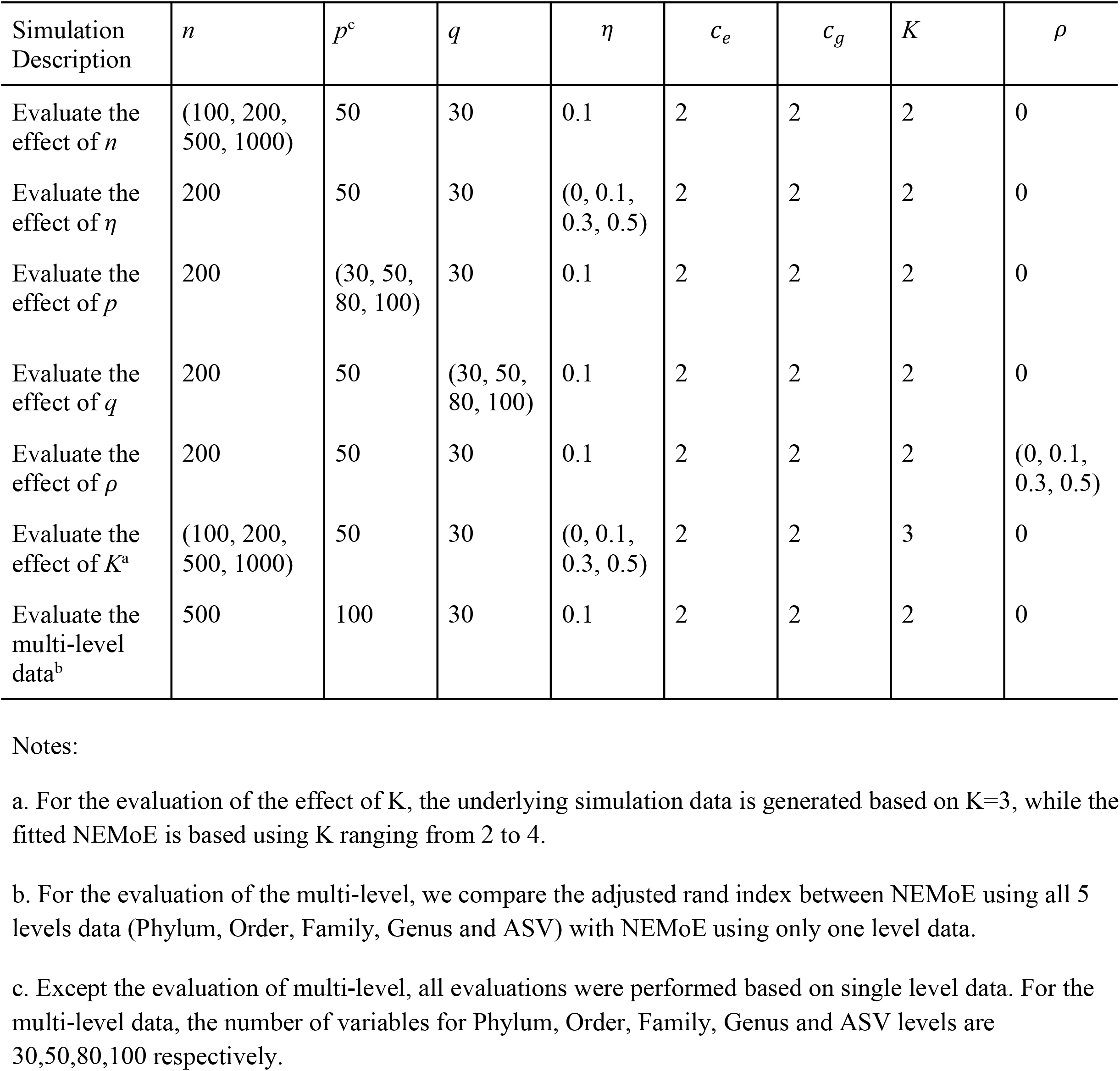
Summary of Simulation settings.

## References

1. Li, H. Microbiome, Metagenomics, and High-Dimensional Compositional Data Analysis. Annu. Rev. Stat. Appl. 2, 73–94 (2015).

2. Wu, G. D. et al. Linking long-term dietary patterns with gut microbial enterotypes. Science 334, 105–108 (2011).

3. Cho, J. H. & Abraham, C. Inflammatory bowel disease genetics: Nod2. Annu. Rev. Med. 58, 401–416 (2007).

4. Pascal, V. et al. A microbial signature for Crohn’s disease. Gut 66, 813–822 (2017).

5. Qin, J. et al. A metagenome-wide association study of gut microbiota in type 2 diabetes. Nature 490, 55–60 (2012).

6. Koeth, R. A. et al. Intestinal microbiota metabolism of L-carnitine, a nutrient in red meat, promotes atherosclerosis. Nat. Med. 19, 576–585 (2013).

7. Lubomski, M. et al. Parkinson’s disease and the gastrointestinal microbiome. J. Neurol. 267, 2507–2523 (2020).

8. Yu, D. et al. Long-term diet quality is associated with gut microbiome diversity and composition among urban Chinese adults. Am. J. Clin. Nutr. 113, 684–694 (2021).

9. Xu, Z. & Knight, R. Dietary effects on human gut microbiome diversity. Br. J. Nutr. 113 Suppl, S1–5 (2015).

10. McBurney, M. I. et al. Establishing What Constitutes a Healthy Human Gut Microbiome: State of the Science, Regulatory Considerations, and Future Directions. J. Nutr. 149, 1882–1895 (2019).

11. De Filippis, F. et al. High-level adherence to a Mediterranean diet beneficially impacts the gut microbiota and associated metabolome. Gut 65, 1812–1821 (2016).

12. Read, M. N. & Holmes, A. J. Towards an Integrative Understanding of Diet–Host–Gut Microbiome Interactions. Front. Immunol. 8, 538 (2017).

13. Holmes, A. J. et al. Diet-Microbiome Interactions in Health Are Controlled by Intestinal Nitrogen Source Constraints. Cell Metab. 25, 140–151 (2017).

14. Cronin, P., Joyce, S. A., O’Toole, P. W. & O’Connor, E. M. Dietary Fibre Modulates the Gut Microbiota. Nutrients 13, (2021).

15. David, L. A. et al. Diet rapidly and reproducibly alters the human gut microbiome. Nature 505, 559–563 (2014).

16. Hegelmaier, T. et al. Interventional Influence of the Intestinal Microbiome Through Dietary Intervention and Bowel Cleansing Might Improve Motor Symptoms in Parkinson’s Disease. Cells 9, (2020).

17. Zeevi, D. et al. Personalized Nutrition by Prediction of Glycemic Responses. Cell 163, 1079–1094 (2015).

18. Asnicar, F. et al. Microbiome connections with host metabolism and habitual diet from 1,098 deeply phenotyped individuals. Nat. Med. 27, 321–332 (2021).

19. Gut microbiota and nutritional profiles of Parkinson’s disease patients. -MDS Abstracts. https://www.mdsabstracts.org/abstract/gut-microbiota-and-nutritional-profiles-of-parkinsons-disease-patients/ (2021).

20. Liang, D. et al. Correction to: Involvement of gut microbiome in human health and disease: brief overview, knowledge gaps and research opportunities. Gut Pathog. 11, 57 (2019).

21. Schulz, C.-A., Oluwagbemigun, K. & Nöthlings, U. Advances in dietary pattern analysis in nutritional epidemiology. Eur. J. Nutr. 60, 4115–4130 (2021).

22. Tebani, A. & Bekri, S. Paving the Way to Precision Nutrition Through Metabolomics. Front Nutr. 6, 41 (2019).

23. Jannasch, F. et al. Exploratory dietary patterns: a systematic review of methods applied in pan-European studies and of validation studies. Br. J. Nutr. 120, 601–611 (2018).

24. Schulze, M. B. et al. Food based dietary patterns and chronic disease prevention. BMJ 361, k2396 (2018).

25. Hughes, R. L. et al. The Role of the Gut Microbiome in Predicting Response to Diet and the Development of Precision Nutrition Models. Part II: Results. Adv. Nutr. 10, 979–998 (2019).

26. Hose, A. J. et al. Excessive Unbalanced Meat Consumption in the First Year of Life Increases Asthma Risk in the PASTURE and LUKAS2 Birth Cohorts. Front. Immunol. 12, 651709 (2021).

27. Tap, J. et al. Diet and gut microbiome interactions of relevance for symptoms in irritable bowel syndrome. Microbiome 9, 74 (2021).

28. Patrick, E. et al. A multi-step classifier addressing cohort heterogeneity improves performance of prognostic biomarkers in three cancer types. Oncotarget 8, 2807–2815 (2017).

29. Tan, A. H. et al. Gut Microbial Ecosystem in Parkinson Disease: New Clinicobiological Insights from Multi-Omics. Ann. Neurol. 89, 546–559 (2021).

30. Yuksel, S. E., Wilson, J. N. & Gader, P. D. Twenty years of mixture of experts. IEEE Trans. Neural Netw. Learn Syst. 23, 1177–1193 (2012).

31. Lê Cao, K.-A., Meugnier, E. & McLachlan, G. J. Integrative mixture of experts to combine clinical factors and gene markers. Bioinformatics 26, 1192–1198 (2010).

32. Huynh, B. T. & Chamroukhi, F. Estimation and Feature Selection in Mixtures of Generalized Linear Experts Models. arXiv preprint at https://arxiv.org/abs/1907.06994.

33. Zou, H. & Hastie, T. Regularization and variable selection via the elastic net. J. R. Stat. Soc. B. 67 301–320 (2005).

34. Costea, P. I. et al. Enterotypes in the landscape of gut microbial community composition. Nat Microbiol 3, 8–16 (2018).

35. Arumugam, M. et al. Enterotypes of the human gut microbiome. Nature 473, 174–180 (2011).

36. Wallen, Z. D. et al. Characterizing dysbiosis of gut microbiome in PD: evidence for overabundance of opportunistic pathogens. NPJ Parkinsons Dis. 6, 11 (2020).

37. Gerhardt, S. & Mohajeri, M. Changes of Colonic Bacterial Composition in Parkinson’s Disease and Other Neurodegenerative Diseases. Nutrients 10 708 (2018).

38. Lubomski, M. et al. The impact of device-assisted therapies on the gut microbiome in Parkinson’s disease. J. Neurol. (2021) doi:10.1007/s00415-021-10657-9.

39. Keshavarzian, A. et al. Colonic bacterial composition in Parkinson’s disease. Mov. Disord. 30, 1351–1360 (2015).

40. Romano, S. et al. Meta-analysis of the Parkinson’s disease gut microbiome suggests alterations linked to intestinal inflammation. NPJ Parkinsons Dis. 7, 1–13 (2021).

41. Callahan, B. J. et al. DADA2: High-resolution sample inference from Illumina amplicon data. Nat. Methods 13, 581–583 (2016).

42. Yilmaz, P. et al. The SILVA and ‘All-species Living Tree Project (LTP)’ taxonomic frameworks. Nucleic Acids Res. 42 D643–D648 (2014).

43. Quast, C. et al. The SILVA ribosomal RNA gene database project: improved data processing and web-based tools. Nucleic Acids Res. 41, D590–6 (2013).

44. Jin, M. et al. Analysis of the Gut Microflora in Patients With Parkinson’s Disease. Front. Neurosci. 13, 1184 (2019).

45. Aho, V. T. E. et al. Relationships of gut microbiota, short-chain fatty acids, inflammation, and the gut barrier in Parkinson’s disease. Mol. Neurodegener. 16, 6 (2021).

46. Kang, Y. et al. Gut Microbiota and Parkinson’s Disease: Implications for Faecal Microbiota Transplantation Therapy. ASN Neuro. 13 175909142110162 (2021).

47. Bullich, C., et al. Gut Vibes in Parkinson’s Disease: The Microbiota-Gut-Brain Axis. Mov. Disord. Clin. Pract. 6, 639–651 (2019).

48. Fernandes, A. D. et al. Unifying the analysis of high-throughput sequencing datasets: characterizing RNA-seq, 16S rRNA gene sequencing and selective growth experiments by compositional data analysis. Microbiome 2, 15 (2014).

## References

49. Palavra, N. C. et al. Increased Added Sugar Consumption Is Common in Parkinson’s Disease. Front. Nutr. 8, 628845 (2021).

50. Hill-Burns, E. M. et al. Parkinson’s disease and Parkinson’s disease medications have distinct signatures of the gut microbiome. Mov. Disord. 32, 739–749 (2017).

51. Aho, V. T. E. et al. Gut microbiota in Parkinson’s disease: Temporal stability and relations to disease progression. EBioMedicine 44, 691–707 (2019).

52. Weis, S. et al. Effect of Parkinson’s disease and related medications on the composition of the fecal bacterial microbiota. NPJ Parkinsons Dis. 5 (2019).

53. Pietrucci, D. et al. Dysbiosis of gut microbiota in a selected population of Parkinson’s patients. Parkinsonism Relat. D. vol. 65 124–130 (2019).

54. Scheperjans, F. et al. Gut microbiota are related to Parkinson’s disease and clinical phenotype. Mov. Disord. 30, 350–358 (2015).

55. Dong, M., Li, L., Chen, M., Kusalik, A. & Xu, W. Predictive analysis methods for human microbiome data with application to Parkinson’s disease. PLoS One 15, e0237779 (2020).

56. Ma, S. et al. Population Structure Discovery in Meta-Analyzed Microbial Communities and Inflammatory Bowel Disease. bioRxiv preprint at https://www.biorxiv.org/content/10.1101/2020.08.31.261214v1.

57. Simpson, S. J. et al. The Geometric Framework for Nutrition as a tool in precision medicine. Nutr. Healthy Aging 4, 217–226 (2017).

58. Raubenheimer, D. & Simpson, S. J. Nutritional Ecology and Human Health. Annu. Rev. Nutr. vol. 36 603–626 (2016).

59. Makkuva, A., Oh, S., Kannan, S. & Viswanath, P. Learning in Gated Neural Networks. in Proceedings of the Twenty Third International Conference on Artificial Intelligence and Statistics (eds. Chiappa, S. & Calandra, R.) vol. 108 3338–3348 (PMLR, 2020).

60. Fruhwirth-Schnatter, S., Celeux, G. & Robert, C. P. Handbook of Mixture Analysis. (CRC Press, 2019).

61. Robert, C. Machine Learning, a Probabilistic Perspective. CHANCE vol. 27 62–63 (2014).

62. Städler, N., Bühlmann, P. & van de Geer, S. 𝓁1-penalization for mixture regression models. TEST 19 209–256 (2010).

63. Khalili, A. & Chen, J. Variable Selection in Finite Mixture of Regression Models. J. Am. Stat. Assoc. 102 1025–1038 (2007).

64. Friedman, J., Hastie, T. & Tibshirani, R. Regularization Paths for Generalized Linear Models via Coordinate Descent. J. Stat. Softw. 33, 1–22 (2010).

65. Chen, J. & Li, H. Variable Selection for Sparse Dirichlet-Multinomial Regression with an Application to Microbiome Analysis. Ann. Appl. Stat. 7, (2013).

66. Deek, R. A. & Li, H. A Zero-Inflated Latent Dirichlet Allocation Model for Microbiome Studies. Front. Genet. 11, 602594 (2020).

67. Wang, T. & Zhao, H. Constructing Predictive Microbial Signatures at Multiple Taxonomic Levels. J. Am. Stat. Assoc. 112 1022–1031 (2017).

